# From Lab to Life: Enhancing Wearable Airbag Fall Detection Performance with Minimal Real-World Data

**DOI:** 10.1101/2025.05.22.25328173

**Authors:** Kyle Embry, Sajjad Daneshgar, Katelyn Aragon, Jonathan Mendley, Kavya Sudhir, Arun Jayaraman

## Abstract

Falls are the leading cause of accidental injury or death among older adults, particularly those with neurological conditions like stroke or Parkinson’s disease (PD) which impair mobility and balance. In these populations, falls are nearly unavoidable, but wearable airbags equipped with pre-impact fall detection algorithms may offer life-saving protection. However, collecting real-world fall data to train these pre-impact algorithms is time-consuming and costly, often leading to the use of simulated falls for model training. This study aimed to 1) identify the best-performing machine learning algorithms for real-world pre-impact fall detection using only simulated falls for training (independent environment approach) and 2) evaluate whether integrating a small amount of real-world data improves detection performance (combined environment approach). Real-world fall data were collected from 22 individuals (N = 12 stroke; N = 10 PD) wearing a waist-mounted wearable airbag device with inertial measurement units (IMUs). A simulated dataset (645 falls, 979 non-falls) was used to train models, while real-world data (32 falls, 32 non-falls) were used for testing and refining models. In the independent environment approach, random forest classifiers achieved the highest performance (F1 = 0.86). Incorporating real-world data and model fine-tuning improved performance, with the best combined environment model reaching an F1 score of 0.93. Feature analysis identified gyroscopic data as the most critical for classification. While real-world data collection remains challenging, integrating even a small amount of real-world falls significantly improves model generalizability. These findings highlight the potential of pre-impact fall detection algorithms for real-world applications, particularly in high-risk populations.

**One Sentence Summary:** Integrating even a small amount of real-world fall data into machine learning models trained on simulated falls significantly improves the performance of pre-impact fall detection algorithms for use in wearable airbags.

## INTRODUCTION

Advancements in medical technology have significantly improved health outcomes, including life expectancy. While this is a positive development, an aging population also presents challenges. One major challenge is the increased prevalence of falls, which are the leading cause of accidental injury in the older adult population in the United States (*1*) and the second leading cause of unintentional injury deaths globally (*2*). The fall rate for individuals aged 65 and over is greater than 25% (*1*), with 37% of falls resulting in injuries that require reduced activity or medical attention (*3*). Aging also leads to greatly increased rates of many neurological disorders like stroke or Parkinson’s disease (*4*, *5*), which further increase the risk of falling. Individuals with neurological conditions fall at rates three times higher than those without such conditions (*6*). Among most common neurological conditions, those with stroke or Parkinson’s disease (PD) were found to have the highest risk of falls (*6*), with a likelihood of falling six and five times greater than those without, respectively, over a one-year period.

The physical and psychological consequences of falls are profound, often resulting in chronic pain, injuries, depression, and social isolation (*7*). Among falls requiring hospitalization, approximately 50% involve hip fractures (*8*). Due to the variation in symptoms and motor deficits, standard interventions such as medications and exercise programs have shown limited effectiveness in preventing falls for people with neurological conditions (*9–11*), urging the development of alternative strategies. Fall protection or detection technologies offer alternative solutions for individuals with high risk of falls. While low-tech options are available, their practicality remains a challenge; for example, hip pads have very low adherence rates, and in-home crash mats can even increase the likelihood of falls due to an unstable surface (*12*, *13*).

More advanced systems include technologies for detecting falls either before or after the impact, also known as pre-impact and post-impact fall detection systems, respectively (*14*). In scenarios where falls cannot be avoided, pre-impact fall detection systems predict falls before they occur (*15*) and can be used with other technologies, such as wearable airbags (*16*) or gyroscopic actuators (*17*), to mitigate fall-related injuries. In contrast, post-impact systems are developed to identify when a fall has already occurred, in order to alert caregivers or emergency services and seek medical attention (*18*), reducing the effects of ‘long-lies’ (*14*, *19*). Examples of these fall detection systems include Context-Aware Systems (CAS) and wearable devices. CAS are typically set-up in one’s home and can monitor the user via pressure sensors, microphones, cameras, radars, ultrasonic or infrared sensors (*20*). Alternatively, wearable devices utilize sensors (inertial measurement units, magnetometers, and electromyography) (*21*) that can be located in numerous places on the body, such as the wrists, waist, chest, and ankles (*22*), and capture whole body motions during a fall.

While previous studies have shown successful fall detection and protection rates, they have tested their systems on general populations, often excluding individuals such as older adults (*23*) and individuals with neurological disorders (*19*, *24–26*), despite being populations with significant fall risks. This is a crucial step for algorithm development, as the performance of the fall detection system is influenced by the data it learns from. For example, Botonis et al., (*27*) developed a stroke-specific model and found increased model performance for detecting falls for individuals with stroke when the training data was also from individuals with stroke instead non-impaired adults.

The importance of the training data presents another key challenge for the development of fall detection algorithms - the environmental context in which the algorithms are developed.

Recording real-world falls requires long periods of continuous monitoring, leading to costly and time-consuming data collection (*28*). Unsurprisingly, most fall detection algorithms are trained on data collected in laboratory settings, where the falls are simulated (i.e., individuals are pushed or instructed to fall on purpose) (*23*, *29*). However, simulated falls differ from real-world falls in acceleration and velocity parameters (*20*, *30*). This limits the accuracy and reliability of detection systems that are developed based on simulated falls, when used in real-world environments (*28*). Due to the high expense of collecting real-world fall data, particularly for people with neurological conditions, training pre-impact fall detection algorithms exclusively from real-world data may not be a feasible solution. Instead, advancements in machine learning may allow fall detection algorithms to be developed using primarily simulated data, with small amounts of real-world data incorporated through transfer learning (*31*) and fine-tuning techniques, which could enhance the algorithm’s generalizability to real-world contexts, while improving the efficiency of the development process.

Therefore, this study aimed to explore the relations between real-world and simulated fall data to 1) determine what type of machine learning algorithm has the best pre-impact fall detection performance (F1 score) on real-world fall data, when trained only on simulated data and 2) develop an integrated machine learning model that utilizes both environments (simulated and real-world) to understand fall patterns across environments and further improve the detection performance (F1 score).

## RESULTS

This study employed a dual-dataset approach, combining real-world fall data with a simulated fall dataset with various methods to validate the performance of 11 different pre-impact fall detection algorithms. Both datasets contain data from individuals with stroke or Parkinson’s disease, who were wearing a wearable airbag device with IMU (inertial measurement unit) sensors at the waist. The device used was a WOLK hip airbag (*32*), which features hip-covering airbags to protect against hip fractures.

The simulated dataset (*27*), included a total of 645 falls and 979 non-falls, which were used to train the pre-impact fall detection algorithms. In the real-world dataset, wearable device activations were recorded for 9 out of 22 participants, containing 32 falls and 18 non-falls (false activations). These participants performed activities that generated an additional 394 non-fall events during in lab visits. To address the fall (N = 32) vs non-fall (N = 412) class imbalance in the real-world dataset, non-fall events were randomly under-sampled to match the number of fall events. In both datasets, three pre-impact windows were used for each fall or non-fall event to extract features, as well as train and test the machine learning algorithms on classifying each pre-impact window as a fall or non-fall event.

The purpose of developing different machine learning algorithms was to determine the best method for classifying real-world fall events. Two comprehensive approaches were designed to use the simulated dataset to train the machine learning algorithms and test on the real-world dataset. In the first approach (independent environment models), the simulated and real-world dataset were used in training and testing the algorithms, respectively. The models in the second approach (combined environment models) used both the simulated dataset and a subset of the real-world dataset in model training and the remainder of the real-world dataset to test the algorithm performance. The performance of all models were measured using F1 score, please see Equations 1-3 for details.

### Independent Environment Models

In the Independent Environment Models, simulated data was used exclusively for training, and real-world data was used only for testing, keeping the two environments independent. Among six tested machine learning models, random forest had the best classification performance on classification of the real-world fall windows (F1 score = 0.86). In classifying simulated fall windows in the cross-validation process, XGBoost had the best performance (Cross-validation F1 score = 0.94). All models showed a higher cross-validation F1 score on the simulated data than the test F1 score on the real-world data. Performance results and best hyperparameters of all independent environment models are listed on Table 1.

**Table 1.** Independent Environment Models performance. The performance of all independent environment machine learning models for classification of fall windows in the real-world environment. Each model is cross-validated on a subset (20%) of the simulated dataset, grouped by participants. A set of selected hyperparameters (names provided from the scikit-learn Python toolbox (*33*)), along with the F1 score, cross-validation F1 score, precision, recall, and AUC (Area Under the Curve) values are listed.

| Model | Algorithm | Hyperparameters | F1 Score | Cross Validation F1 score | Precision | Recall | AUC |
| --- | --- | --- | --- | --- | --- | --- | --- |
| 1 | K-nearest neighbor | Metric = 'manhattan'<br>N_neighbors = 9<br>Weights = distance | 0.71 | 0.82 | 1.00 | 0.55 | 0.90 |
| 2 | Decision Tree | Criterion = 'gini'<br>Max_depth = 10<br>Max_features = 'sqrt'<br>Min_samples_leaf = 4<br>Min_samples_split = 5<br>Splitter = 'best' | 0.82 | 0.91 | 0.95 | 0.72 | 0.90 |
| 3 | AdaBoost | Algorithm = 'SAMME'<br>Learning_rate = 0.3<br>N_estimators = 200 | 0.83 | 0.92 | 1.00 | 0.71 | 0.98 |
| 4 | Logistic Regression | C = 1<br>Max_iter = 100<br>Penalty = 'l2'<br>Solver = 'liblinear' | 0.83 | 0.93 | 0.99 | 0.72 | 0.99 |
| 5 | XGBoost | N_estimators = 200<br>Max_depth = 5<br>Learning_rate = 0.1<br>Subsample = 0.8<br>Colsample_bytree = 1<br>Gamma = 0 | 0.85 | 0.94 | 1.00 | 0.73 | 0.99 |
| 6 | Random Forest | N_estimators = 200<br>Max_features = 'sqrt'<br>Max_depth = 10<br>Min_samples_split = 5<br>Min_samples_leaf = 4 | 0.86 | 0.93 | 1.00 | 0.74 | 0.99 |

In the best-performing model, random forest (Table 1; Model 6), feature contributions to the overall model performance were evaluated separately for the training and test datasets. For the simulated training data, the random forest’s feature importance algorithm ranked all features based on their classification relevance. The top eight features were all derived from gyroscope signals, representing various characteristics of angular velocity (Fig. 1). In the real-world test dataset, the impact of each feature was assessed using a permutation technique. Feature values were randomly shuffled over thirty iterations and the change in the model’s F1 score after every permutation was recorded, averaged and their absolute values were taken to quantify the importance of each feature in the test set. Important features, when their data is shuffled in this manner, will change the F1 score more.

**Fig. 1.**
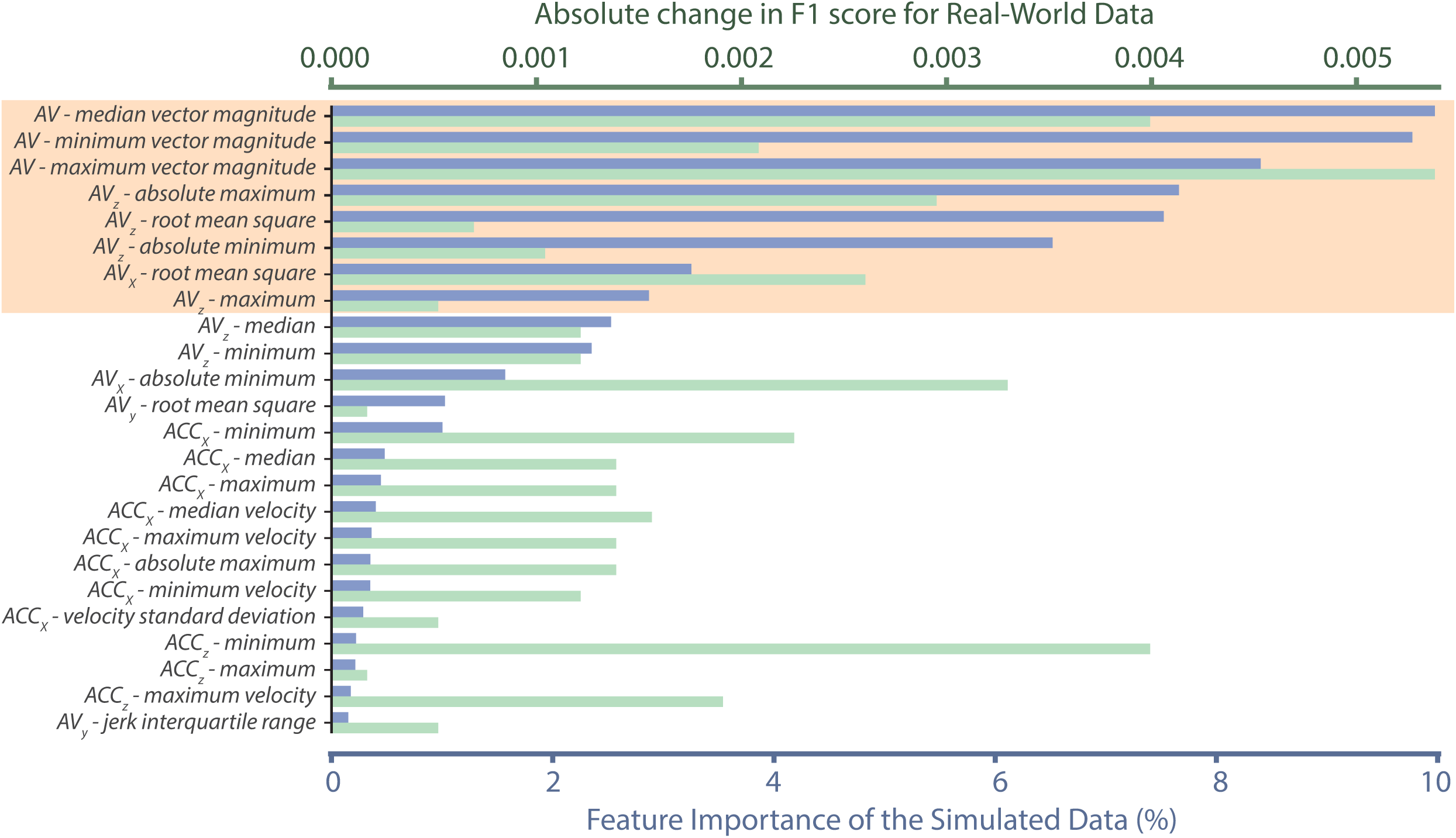
Most important features in the simulated and real-world fall data. The most important features in the simulated fall data are shown in blue bars, with the top eight important features highlighted in orange. For each feature, values in the test dataset were permuted, and the resulting absolute changes in F1 scores are shown in green bars. Features with no F1 score change after permutation were excluded in the visualization. AV = angular velocity; ACC = acceleration.

This analysis revealed that the most influential features in the training set (simulated data), also had the greatest influence in the test set (real-world data), with their permutations causing the largest changes in F1 score (Fig. 1). This indicates a good model generalization, consistent with the cross-validation and test F1 scores of 0.93 and 0.86 (Table 1; Model 6). Although some of the less important features in the trained model–particularly from acceleration signals–became important in the real-world environment, their permutation scores remained smaller and evenly distributed across features (Fig. 1). Unlike the differences in important features across fall and non-fall classes, the distribution of these features across environments remained similar (Fig. S1 and S2). Overall, these results indicate that both environments largely agree about the most important features, but differ somewhat on their rankings, and disagree about many of the less important features.

### Combined Environment Models

The combined environment models were trained using data from both simulated and real-world environments, using the same random forest design as in Table 1, Model 6. In the fine-tuned and weighted learning models, half of the real-world data, grouped by subjects, was oversampled and added to the training process. The same steps were completed for the transfer learning models, but without any oversampling. The classification performance of all models were evaluated on the remaining unseen subset of the real-world data. Performance results of all combined environment models are listed in Table 2. Most combined environment models outperformed the independent models (Table 1). The best performance was obtained using the fine-tuned random forest, with a F1 score of 0.93, improving by 0.07 and matching the cross-validation F1 score of the simulated data when comparing to the independent random forest model (Table 2; Model 6).

**Table 2.** Combined Environment Models performance. The performance of all combined environment machine learning models for classification of fall windows in the real-world environment. The first model listed in the table is the reference of the best-performing independent environment model in Table 1 (Model 6). For each model, the split strategy, along with the F1 score, precision, recall and AUC values are listed.

| Model | Algorithm | Split Strategy | F1 Score | Precision | Recall | AUC |
| --- | --- | --- | --- | --- | --- | --- |
| 6 | Random Forest<br>(Reference; Table 1) | Training set: all simulated data<br>Test set: all real-world data | 0.86 | 1.00 | 0.74 | 0.99 |
| 7 | SER | Training set: all simulated data<br>Training set for refining: 5 real-world subjects<br>Test set: 4 remaining real world subjects | 0.80 | 0.99 | 0.69 | 0.84 |
| 8 | Weighted Learning<br>Random Forest | Training set: all simulated data<br>Training set for refining: 5 real-world subjects with oversampled data<br>Test set: 4 remaining real-world subjects with oversampled data | 0.85 | 0.94 | 0.78 | 0.86 |
| 9 | MIX (SER + STRUT) | Same as Model 7 | 0.88 | 0.99 | 0.79 | 0.89 |
| 10 | STRUT | Same as Model 7 | 0.90 | 0.94 | 0.87 | 0.91 |
| 11 | Fine-Tuned Random<br>Forest | Same as Model 8 | 0.93 | 0.91 | 0.95 | 0.92 |

## DISCUSSION

In this study, we evaluated the performance of pre-impact fall detection algorithms in distinguishing falls from non-falls among individuals with neurological diseases, using data from both laboratory and real-world environments. Our results demonstrate that the model type used can have a meaningful effect on how well a classifier will generalize from a simulated to real-world environment, with random forests performing the best of the model types tested (Table 1, Model 6). Additionally, fine-tuning techniques and integrating a portion of real-world data into the training process further improved classification performance (F1 = 0.93; Table 2, Model 11).

The observed 0.07 increase in the F1 score suggests differences between simulated and real-world falls in individuals with neurological diseases, which we will explore in the following sections.

### Fall vs. Non-Fall Classification

Our classification models successfully distinguished falls from non-fall windows. When considering real-world and simulated settings as independent environments in the machine learning algorithms, the random forest algorithm can classify real-world falls from non-falls with an F1 score of 0.86 (Table 1; Model 6). This performance was achieved when the model was trained on only the simulated dataset. Combining environments into the training set increased the F1 score to 0.93 (Table 2; Model 11).

This study specifically classified rear or hip-impacting falls as “falls”, whereas forward falls were categorized as “non-falls”. Although forward falls are among the most common fall directions (*34*), they are often mitigated by reflexive responses, with the knees and hands absorbing much of the impact (*35*). Defining rear and hip-impacting falls as “falls” also aligns with the design on the WOLK wearable device used in this study, which features airbags on the rear and sides for hip protection. Limiting “falls” to only rear or hip impacting falls and excluding forward falls may reduce the generalizability of the machine learning models in recognizing diverse fall patterns; however, it enhances wearable device’s specificity by excluding non-hip-impact forward falls and focusing on the types of falls most relevant to its design and intended protective function. The method used in this study could be adapted to classify other types of falls as “falls”, if using other developed wearables to protect other body regions, such as the knees of head.

A significant challenge in real-world experiments is the data imbalance between the simulated and real-world events, caused by the lower acquisition rate of real falls (*36*, *37*) and the difficulty of capturing a range of activities of daily living (ADLs) due to their unpredictable and often unstructured nature. To address this challenge and enhance the representation of real-world events in the dataset, we incorporated a combination of in-laboratory ADLs and real-world non-fall events. While this approach introduces some limitations, previous studies have not identified significant biomechanical differences in ADL patterns between laboratory and real-world environments (*38*, *39*). This suggests that in-lab ADLs can serve as reasonable approximations of real-world ADLs, and help to balance the dataset.

In contrast to post-impact algorithms and systems (*40*, *41*) which notify caregivers only after a fall has occurred, our pre-impact, single-sensor detection algorithm utilized multiple pre-impact windows with different lead times (*27*, *42*). This approach enables real-time intervention and practicality for daily use and makes it suitable for integration into wearable devices and fall- injury mitigation.

### Simulated vs. Real-World Falls

In this study, we explored different approaches for utilizing simulated fall data and limited amounts of real-world data to develop high-performance pre-impact fall detection algorithms, as measured by the F1 score. When trained exclusively on simulated data, the algorithms showed a drop in performance when detecting real-world falls (F1 score = 0.86; Table 1, Model 6) compared to their performance on simulated falls (Cross-validation F1 score = 0.93; Table 1, Model 6). However, incorporating both simulated and real-world data during training improved real-world fall detection, with the best performing model achieving an F1 score comparable to the cross-validation score (F1 score = 0.93; Table 2, Model 11), which differs from previous studies (*20*, *28*, *30*, *43*). While the improvement in F1 score from 0.86 to 0.93 may seem small, the recall rate increased from 0.74 to 0.95. Put another way, this improvement equates to reducing missed true falls from 26% to 5%--a nearly five-fold decrease.

Data collected from simulated falls differ from real-world falls for a variety of reasons, including changes in reflex responses when falls are anticipated, and the speed and intensity of falls that are possible to safely perform in a laboratory setting (*44*). Due to these differences in movement, the sensor readings during the pre-impact, impact, and post-impact phases of a fall also differ in terms of acceleration and angular velocity (*28*). These differences contribute to why algorithms trained on simulated data tend to perform worse on real-world data (*28*, *30*). To address this, we examined how the combined environment methods (Table 2) could adapt to both environments and improve real-world fall detection performance. Fine-tuning and transfer learning models showed improved performance, with F1 scores of 0.93 and 0.90, respectively (Table 2, Model 11 and 10). This improvement suggests that differences in fall patterns across environments can be compensated for using fine-tuned random forests and transfer learning models. One of the key factors in our fall detection models was the role of gyroscopic features, which emerged as the most critical features in distinguishing falls from non-falls in both environments (Fig. 1). These features–including the magnitude, absolute value and vector magnitude of angular velocity–were highly influential in model performance. While previous studies have explored gyroscopic features in both simulated (*45*) and real-world falls (*46*), most research has relied on acceleration signals (*30*, *39*, *47–50*), likely due to their lower processing demands and more straightforward representation of fall motion. However, our findings demonstrated the ability of gyroscopic features to classify falls, as the models that generalized the best from simulated to real-world data relied heavily on gyroscopic features.

Although gyroscopic features can generally classify fall and non-fall events, the discrepancy in feature importance and absolute permutation scores (Fig. 1) highlights differences in simulated and real-world data (Table 1). While gyroscopic features and their interactions dominated prediction power in the simulated environment, real-world fall and non-fall patterns showed a more balanced feature importance distribution across gyroscopic and acceleration features (Fig. 1). Previous studies (*28*, *30*) have also noted the importance of acceleration features in both environments. However, our findings indicated that individual acceleration features had minimal impact on prediction performance (as reflected in absolute change in F1 scores in Fig. 1), and overall feature distributions did not differ significantly between environments (Fig. S1 and S2). This suggests that specific combinations of features, such as gyroscopic-acceleration interactions, influence the real-world patterns more strongly than the simulated patterns, though no single feature dominates. Although subtle variations across multiple features collectively contribute to a potential meaningful difference between real-world and simulated conditions, our results indicate that combined environment approaches can effectively capture and address the differences between environments, improving the generalizability of pre-impact fall detection algorithms while using only a small amount of real-world data.

Our study suggests that combining both simulated and real-world data is important for improving real-world performance. The oversampling process in the combined environment models helped address data imbalance; however, because oversampling does not introduce new information, real-time evaluation across a broader range of daily activities and a larger sample may provide more reliable classification performance. In this study we combined individuals with stroke or PD, but future work should determine if separating those groups leads to better results. Additionally, the inclusion of ADLs completed in the laboratory, while justified, may still not fully replicate the variability of real-world motion. Future research should aim to capture a broader range of natural ADLs and higher number of falls to enhance the validity of the dataset.

## MATERIALS AND METHODS

### Study Design

This study employed data from two independent environments (simulated in-laboratory data, and real-world data) to evaluate the performance of pre-impact fall detection algorithms in distinguishing falls from non-falls among individuals with stroke or Parkinson’s disease in the real world. The simulated dataset (*27*) was collected from 39 research participants (N = 20 stroke; N = 19 PD), (645 falls and 970 non-falls), under controlled laboratory conditions, and was used to train all machine learning algorithms. For the real-world data collection in this study, a separate set of research participants with stroke or PD (N = 22) were recruited based on inclusion criteria such as age (18–85), prior fall history, and ability to wear a waist-mounted airbag device. Real-world fall data was recorded during daily activities over a six month-period and used to either test or refine the classification models. The algorithms utilized features extracted from pre-impact windows driven from IMU sensor data, with target labels categorized into either fall or non-fall. Model performance was evaluated using the F1 score of the fall class to identify the best classifiers for wearable airbag deployment in real-world scenarios. Signal processing, feature extractions, and all machine learning analyses were performed on Python (3.10.11).

### Participants

In this study, we recruited individuals with chronic stroke (at least six months post diagnosis; N = 12; 64 ± 14; 7 female) or Parkinson’s disease (N = 10; average age ± standard deviation of 72.4 ± 3.4 years; 4 female) for the real-world data collection. Eligibility criteria required research participants to be aged 18-85, report at least on fall within the past six months, and be homebound or community ambulators with a waist circumference of 84-125 cm (to accommodate the wearable airbag device sizing). Exclusion criteria included pregnancy, wheelchair dependence for both indoor and outdoor mobility, and color blindness that could interfere with perceiving the device’s light signals. All participants provided informed consent prior to the study, in compliance with Northwestern University IRB guidelines (NU IRB, IL, USA; STU00209246). This clinical trial study is registered on ClinicalTrials.gov (NCT05076565).

### Device

The WOLK Hip Airbag (Wolk De Heupairbag; Wolk Company, Netherlands) was used to collect motion data (Fig. 2). The WOLK airbag is a wearable device that utilizes three fixed triaxial inertial measurement units (IMU) located at the hips and lower back (aligned with the L3 vertebrae) and a computational unit equipped with the pre-impact fall detection algorithm to detect a fall. These IMUs include accelerometer and gyroscope sensors that sample motion data at 500 Hz. In the event of a fall, the device deploys airbags on each hip using embedded CO2 cartridges. For continuous data recording in the laboratory, a modified WOLK device was utilized to keep the airbags inactive.

**Fig. 2.**
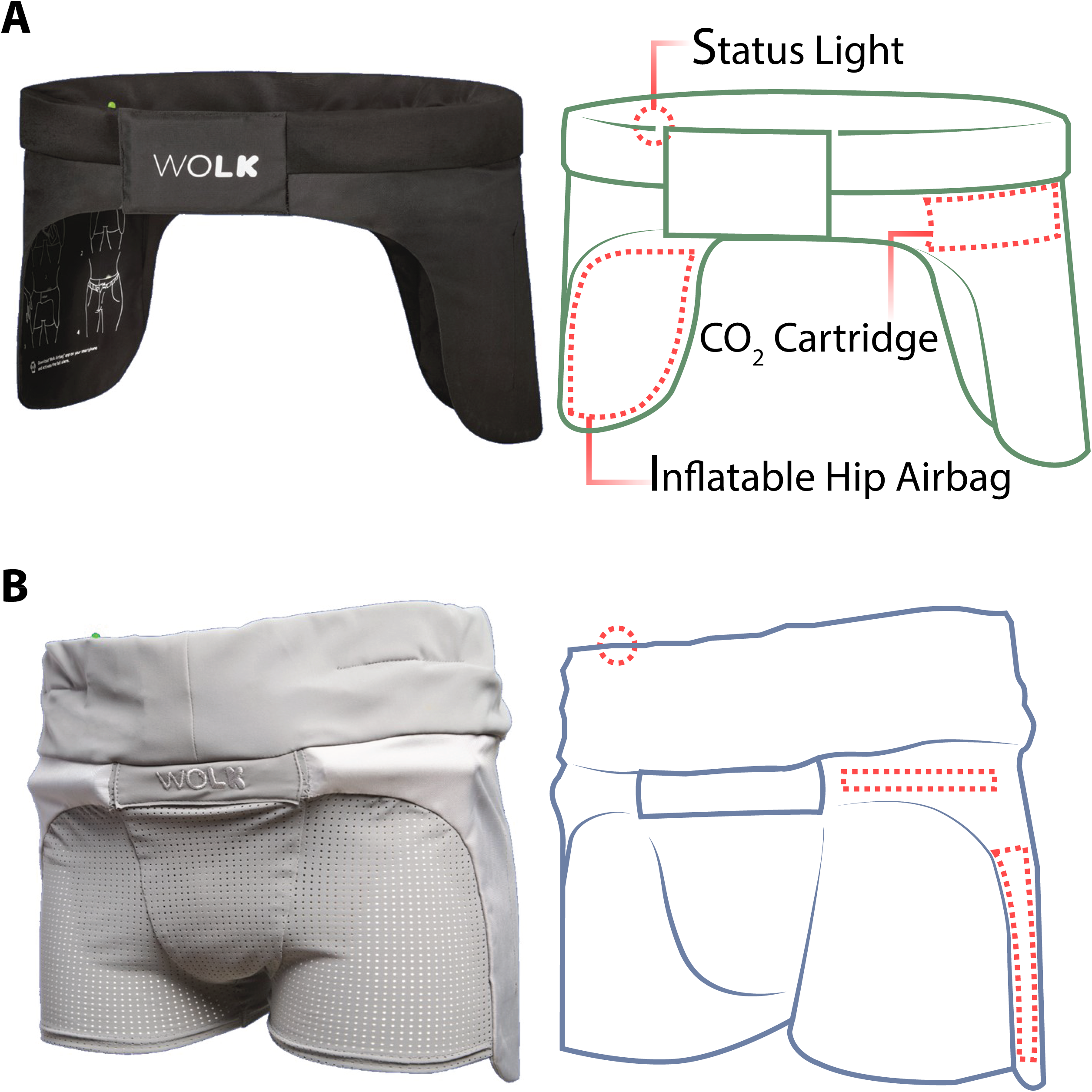
The WOLK Active Airbag System. This device is used in two models **(A** & **B)**. The actual product image is provided by the WOLK (*32*). The belt (top) and short (bottom) are both equipped with deployable hip mounted airbags to protect the hips when a fall is detected.

### Experimental Design

#### Simulated Fall Experiment

The simulated fall dataset was collected in a single laboratory visit (*27*) and included individuals with chronic stroke or PD, and age ranged from 18 to 85. In this study, each research participant completed a series of simulated fall and non-fall tasks (Fig. 3A) while wearing protective equipment (e.g., helmet, neck/knee/elbow guards, padded shorts). This dataset was used to train the fall-prediction algorithms. For the purpose of this study, which focuses on a wearable hip protection system, falls were defined as events where the participant’s hip or rear directly and unintentionally impacted the ground. Non-hip/rear impacting falls, activities of daily living (ADL), and near-fall attempts were all categorized as non-fall events, as there would be no utility in deploying the airbags in these cases. Moreover, forward falls in the simulated dataset, in which the stomach directly impacted the ground, were excluded.

**Fig. 3.**
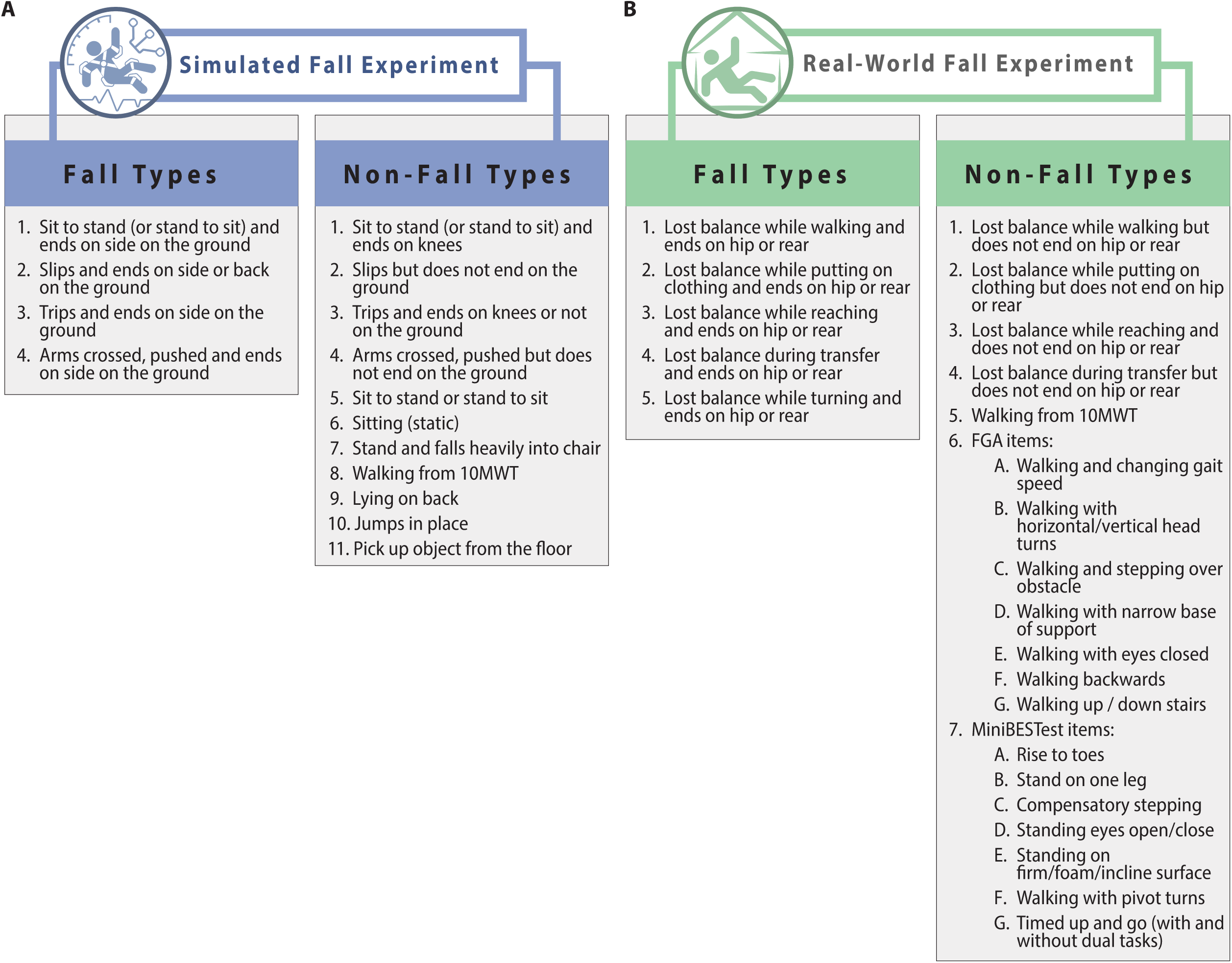
Fall and non-fall events. List of all events completed by research participants in the simulated (**A**) and real-world (**B**) environments. For the real-world research participants, a set of simulated non-fall activities, including the 10 Meter Walk Test (10MWT), Functional Gait Assessment (FGA), and the Mini Balance Evaluation Systems Test (MiniBESTest) were recorded and combined with their real-world non-fall events.

#### Real-World Fall Experiment

Data were collected from research participants over six months in a community setting (Fig. 3B), with research participants attending three laboratory visits: at enrollment, three months and six months post-enrollment. During each visit, research participants were asked to perform physical assessments, including the 10-Meter Walk Test (10MWT), Functional Gait Assessment (FGA), and Mini Balance Evaluation Systems Test (Mini-BESTest). These assessments reflect functional mobility and postural stability, providing representative samples of activities of daily living (ADLs). To address the difficulty of recording and marking non-fall activities in real-world settings, ADLs from the laboratory visits (10MWT, FGA, Mini-BESTest) were added to the non-fall dataset to improve its variability.

Research participants were trained to use the WOLK airbag device at home and then given the device to wear during their daily lives as much as possible. Each WOLK belt was equipped with the WOLK Company’s pre-impact fall detection algorithm (*51*). In the case that a WOLK device detected a fall, IMU data from approximately 6 seconds before and 2 seconds after the fall were recorded and wirelessly transmitted to the research team. Our research team made bi- weekly phone calls to track any incidents of fall, airbag deployments, and obtain more detailed information for each event (e.g., part of body impacted, location, time, activity leading up to fall/deployment, injuries, medical care needed, etc.).

#### Data Processing

To align with most fall detection algorithms that rely on data from a single IMU (*52–56*), data recorded by the two lateral IMUs was discarded, and only the data from the center IMU was retained. Accelerometer signals were band-pass Butterworth filtered (0.1 - 50 Hz). For all fall events, the impact time was defined as the peak acceleration magnitude (Fig. 4A). For non-fall events, which lack an impact time, the peak acceleration magnitude served as a reference to extract similar windows of data (Fig. 4B). From both event types, three pre-impact time windows were marked at 300-200 ms, 250-150 ms, and 200–100 ms before the maximum point (Fig. 4A). For each extracted window, acceleration and gyroscope signals were used to calculate a total of 160 features (Fig. 4C).

**Fig. 4.**
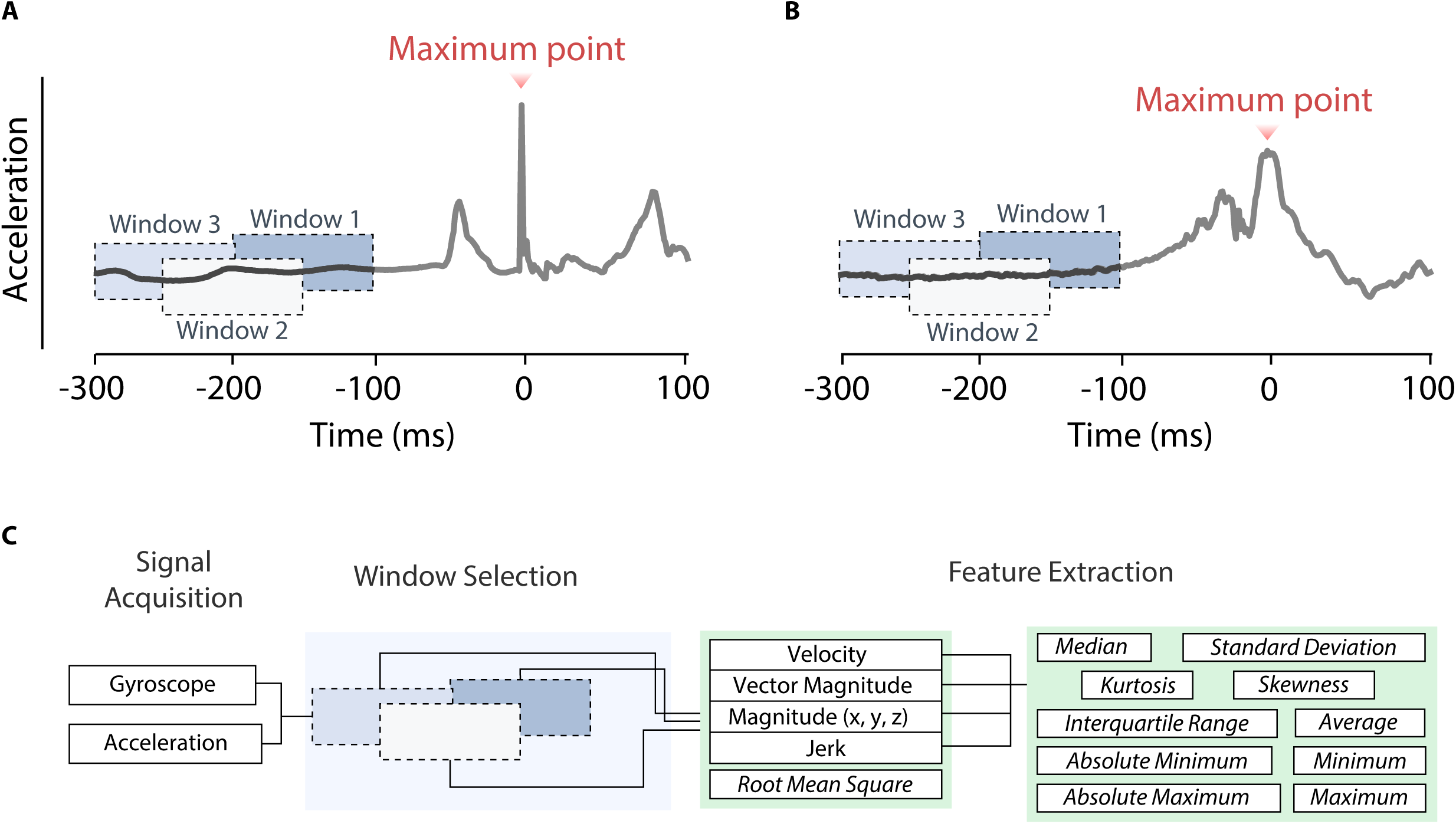
Inertial Measurement Unit feature extraction. Two examples of acceleration signals recorded right during a fall incident (**A**) and a sit to stand task (**B**). For the fall signal, the impact time was marked at the maximum acceleration magnitude. In the non-fall signal, the activity was timestamped and the local maximum acceleration magnitude within the event window was defined as the maximum reference point. In both event types, gyroscope and acceleration signals were extracted, three windows with different lead times were defined, and a set of features were calculated separately for each window (**C**).

Features from the simulated data were used to train the classification algorithms and classify the fall labels in the real-world data. The addition of ADLs from in-lab visits to the real-world non-fall repository resulted in an imbalance between classes (Non-fall events = 412, fall events = 32). To address this issue, a stratified random under-sampling was grouped by participants and applied to the non-fall events. This led to 32 under-sampled non-fall events, distributed as evenly as possible across participants.

### Machine Learning Classification Models

#### Independent Environment Models

The six classification models– 1) K-Nearest Neighbor, 2) Decision Tree, 3) AdaBoost, 4) Logistic Regression, 5) XGBoost, 6) Random Forest–were used to classify fall and non-fall windows. Acceleration and gyroscopic features from the simulated dataset were used to train the algorithms. The real-world data were reserved for only testing the classification algorithms.

Cross-validation was performed using a five-fold split, grouped by participants and class-weighted with hyperparameters tuned using GridSearchCV. Model performance was evaluated using F1 score for the fall class and metrics of cross-validation F1 score, precision, recall, and Area Under the Curve (AUC) were added for the reference. Precision, Recall, and F1 score were defined in Eqs. 1-3.

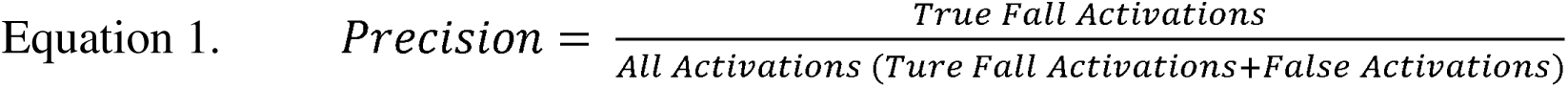

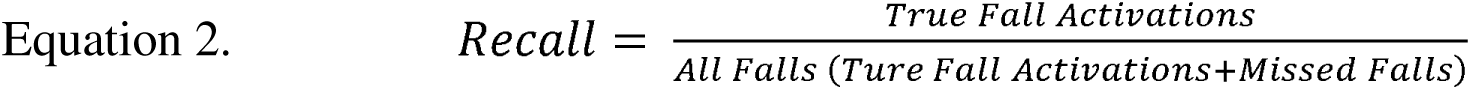

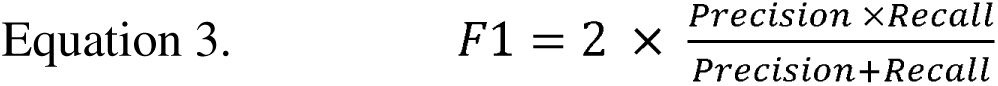

Feature importance was assessed for the best-performing machine learning model (highest F1 score). The selection process was confined to the training and testing data separately to prevent data leakage. In the training (simulated) dataset, feature importance was determined using the model’s built-in ranking algorithm, which evaluates features based on their contribution to classification performance on the validation set. The most relevant features were identified based on this ranking. To assess feature importance in the test (real-world) dataset, a permutation-based importance analysis was applied (*57*). Feature values were randomly shuffled over 30 iterations, and the average absolute change in the model’s F1 score was recorded to quantify each feature’s contribution. The distribution of important features of fall and non-fall classes across simulated and real-world environments were visualized in Fig. S1 and S2.

### Combined Environment Models

These models aimed to further improve generalization across the two environments. The combined environment models were used to adapt the training model with a limited subset of the real-world dataset before final testing on real-world events. In this approach, the best-performing algorithm and its hyperparameters from the independent model environment were used in training the models (Model 6).

### Transfer learning (Models 7, 9, 10)

Transfer learning techniques were applied with the goal of using real-world data to retrain models initially trained with simulated. Three methods– 1) structure expansion/reduction (SER), 2) structure transfer (STRUT), and 3) MIX (SER and STRUT) as described in (*58*, *59*)--were tested. In all transfer learning models, the model (Model 6) was first trained on the “source” data (simulated dataset). The model was then adapted using approximately half of the target data (real-world dataset), grouped by subject, and tested on the remaining half of the target data. During the validation process in the re-training phase, hyperparameters across trees were re- tuned partially (SER) or completely (STRUT). To capture the total variability, all possible combinations were run and the average performance of each model was reported (F1 score, precision, recall, and AUC for the fall class).

### Fine-tuning (Model 11)

Like transfer learning, fine-tuning was applied to improve the model trained on the simulated dataset for better fall classification in the real-world dataset. The best-performing model from the independent environment approach (Model 6) was first trained on the full simulated dataset with fixed hyperparameters, and then re-trained on half of the real-world dataset, grouped by subjects. Unlike transfer learning, the hyperparameters in this approach remained unchanged. During fine-tuning, the weight of features were updated by training on a subset of the real-world dataset. To increase the sample size of the fine-tuning subset, it was augmented using oversampling techniques (Adaptive Synthetic Sampling (ADASYN) and Synthetic Minority Oversampling Techniques (SMOTE)). SMOTE generates new synthetic non-fall samples by interpolating non-fall windows. Additionally, ADASYN was applied to the fall samples to create more synthetic samples in regions where the model struggled. Both fall and non-fall classes were oversampled to have a total of 150 fine-tuning samples in each class. The model was then tested on the remaining real-world data and the same performance metrics were reported.

### Weighted-learning (Model 8)

The weighted-learning model (*60*) was utilized to address the data imbalance between real-world and simulated datasets. The real-world dataset was split into two equal-size groups of subjects: one group for training and the other for testing. To improve the representation of real-world data, the training subset of the real-world data was oversampled using SMOTE. The best-performing algorithm from the independent environment approach was then trained on a combination of the simulated data and an oversampled subset of the real-world data. In the training phase, real-world samples gained extra weights to prioritize their patterns. The model was finally evaluated on the remaining test set using the same performance metrics.

## Data Availability

The data used in the analysis are subject to a Material Transfer Agreement (MTA) and can be made available upon reasonable request to the corresponding author, in accordance with institutional and regulatory guidelines.

## LIST OF SUPPLEMENTARY MATERIALS

Fig. S1 and S2

## ACKNOLEDGEMENT

We sincerely thank all the participants for their time and effort in contributing to this research. Their willingness to take part in this study has been invaluable. We also extend our appreciation to Jeroen van der Heijden from Wolk Company for the hardware and firmware support, which greatly facilitated this work.

## AUTHOR CONTRIBUTIONS

Conceptualization: KE, SD, KA, JM, KS, AJ

Methodology: KE, SD, KA, JM, KS

Investigation: KE, KA, JM, KS

Visualization: SD

Funding acquisition: AJ

Project administration: KE, SD

Supervision: KE, AJ

Writing – original draft: KE, SD, KA, AJ

## DATA AND MATERIALS AVAILABILITY

The code used in this study will be made available online at GitHub upon publication. The data used in the analysis are subject to a Material Transfer Agreement (MTA) and can be made available upon reasonable request to the corresponding author, in accordance with institutional and regulatory guidelines.

---

**Fig. S1. Distribution of the most important features across both environments (Density plot).** Density plots of all features with a nonzero change in the test set (Permutation score). The distributions of simulated (blue) and real-world (green) dataset are grouped by their fall and non-fall class labels, and normalized using kernel density estimates (KDEs), so the areas under the curves are equal. From top to bottom, and left to right, features are sorted by their importance in the training set (simulated data), with the top eight most important features highlighted in orange. AV = angular velocity; ACC = acceleration.

**Fig. S2. Distribution of the most important features across both environments (Box plot).** Box plots of all features with a non-zero F1 score reduction in the test set (Permutation score). The distributions of simulated (blue) and real-world (green) dataset are grouped by their fall and non-fall class labels. Each box, red lines, and error bars represent interquartile range, median, and extreme range of each distribution, respectively. AV = angular velocity; ACC = acceleration.

## REFERENCES

1. Centers for Disease Control and Prevention, National Center for Health Statistics. National Vital Statistics System, Mortality 1999–2021, CDC WONDER Online Database, 2023. Available at: https://wonder.cdc.gov/ucd-icd10.html Accessed February 2025.

2. World Health Organization, Falls – Fact Sheet, WHO, 2021. Available at: https://www.who.int/news-room/fact-sheets/detail/falls Accessed February 2025.

3. B. Moreland, R. Kakara, A. Henry, Trends in Nonfatal Falls and Fall-Related Injuries Among Adults Aged ≥65 Years — United States, 2012–2018. MMWR Morb Mortal Wkly Rep 69, 875–881 (2020).

4. L. Hirsch, N. Jette, A. Frolkis, T. Steeves, T. Pringsheim, The Incidence of Parkinson’s Disease: A Systematic Review and Meta-Analysis. Neuroepidemiology 46, 292–300 (2016).

5. K. Tan, M. Tan, Stroke and Falls—Clash of the Two Titans in Geriatrics. Geriatrics 1, 31 (2016).

6. B. Homann, A. Plaschg, M. Grundner, A. Haubenhofer, T. Griedl, G. Ivanic, E. Hofer, F. Fazekas, C. N. Homann, The impact of neurological disorders on the risk for falls in the community dwelling elderly: a case-controlled study. BMJ Open 3, e003367 (2013).

7. R. Vaishya, A. Vaish, Falls in Older Adults are Serious. Indian J Orthop 54, 69–74 (2020).

8. C.-S. Rau, T.-S. Lin, S.-C. Wu, J. C.-S. Yang, S.-Y. Hsu, T.-Y. Cho, C.-H. Hsieh, Geriatric hospitalizations in fall-related injuries. Scand J Trauma Resusc Emerg Med 22, 63 (2014).

9. N. E. Allen, C. G. Canning, L. R. S. Almeida, B. R. Bloem, S. H. Keus, N. Löfgren, A. Nieuwboer, G. S. Verheyden, T. P. Yamato, C. Sherrington, Interventions for preventing falls in Parkinson’s disease. Cochrane Database of Systematic Reviews 2022 (2022).

10. S. Denissen, W. Staring, D. Kunkel, R. M. Pickering, S. Lennon, A. C. Geurts, V. Weerdesteyn, G. S. Verheyden, Interventions for preventing falls in people after stroke. Cochrane Database of Systematic Reviews 2019 (2019).

11. N. O’Malley, A. M. Clifford, M. Conneely, B. Casey, S. Coote, Effectiveness of interventions to prevent falls for people with multiple sclerosis, Parkinson’s disease and stroke: an umbrella review. BMC Neurol 21, 378 (2021).

12. N. Santesso, A. Carrasco-Labra, R. Brignardello-Petersen, Hip protectors for preventing hip fractures in older people. Cochrane Database of Systematic Reviews 2014 (2014).

13. A. K. Doig, J. M. Morse, The Hazards of Using Floor Mats as a Fall Protection Device at the Bedside. J Patient Saf 6, 68–75 (2010).

14. J. Liu, X. Li, S. Huang, R. Chao, Z. Cao, S. Wang, A. Wang, L. Liu, A review of wearable sensors based fall-related recognition systems. Eng Appl Artif Intell 121, 105993 (2023).

15. N. Otanasap, “Pre-Impact Fall Detection Based on Wearable Device Using Dynamic Threshold Model” in 2016 17th International Conference on Parallel and Distributed Computing, Applications and Technologies (PDCAT) (IEEE, 2016), pp. 362–365.

16. T. Tamura, T. Yoshimura, M. Sekine, M. Uchida, O. Tanaka, A Wearable Airbag to Prevent Fall Injuries. IEEE Transactions on Information Technology in Biomedicine 13, 910–914 (2009).

17. B. T. Sterke, K. L. Poggensee, G. M. Ribbers, D. Lemus, H. Vallery, Light-Weight Wearable Gyroscopic Actuators Can Modulate Balance Performance and Gait Characteristics: A Proof-of-Concept Study. Healthcare 11, 2841 (2023).

18. H. Nguyen, F. Mirza, M. A. Naeem, M. M. Baig, Falls management framework for supporting an independent lifestyle for older adults: a systematic review. Aging Clin Exp Res 30, 1275–1286 (2018).

19. X. Yu, B. Koo, J. Jang, Y. Kim, S. Xiong, A comprehensive comparison of accuracy and practicality of different types of algorithms for pre-impact fall detection using both young and old adults. Measurement 201, 111785 (2022).

20. E. Casilari, C. A. Silva, An analytical comparison of datasets of Real-World and simulated falls intended for the evaluation of wearable fall alerting systems. Measurement 202, 111843 (2022).

21. T. Xu, Y. Zhou, J. Zhu, New Advances and Challenges of Fall Detection Systems: A Survey. Applied Sciences 8, 418 (2018).

22. R. Rucco, A. Sorriso, M. Liparoti, G. Ferraioli, P. Sorrentino, M. Ambrosanio, F. Baselice, Type and Location of Wearable Sensors for Monitoring Falls during Static and Dynamic Tasks in Healthy Elderly: A Review. Sensors 18, 1613 (2018).

23. S. Chaudhuri, H. Thompson, G. Demiris, Fall Detection Devices and Their Use With Older Adults. Journal of Geriatric Physical Therapy 37, 178–196 (2014).

24. S. K. Gharghan, H. A. Hashim, A comprehensive review of elderly fall detection using wireless communication and artificial intelligence techniques. Measurement 226, 114186 (2024).

25. C. Taramasco, T. Rodenas, F. Martinez, P. Fuentes, R. Munoz, R. Olivares, V. H. C. De Albuquerque, J. Demongeot, A Novel Monitoring System for Fall Detection in Older People. IEEE Access 6, 43563–43574 (2018).

26. F. Kausar, M. Mesbah, W. Iqbal, A. Ahmad, I. Sayyed, Fall Detection in the Elderly using Different Machine Learning Algorithms with Optimal Window Size. Mobile Networks and Applications 29, 413–423 (2024).

27. O. K. Botonis, Y. Harari, K. R. Embry, C. K. Mummidisetty, D. Riopelle, M. Giffhorn, M. V. Albert, V. Heike, A. Jayaraman, Wearable airbag technology and machine learned models to mitigate falls after stroke. J Neuroeng Rehabil 19, 1–14 (2022).

28. F. Bagalà, C. Becker, A. Cappello, L. Chiari, K. Aminian, J. M. Hausdorff, W. Zijlstra, J. Klenk, Evaluation of Accelerometer-Based Fall Detection Algorithms on Real-World Falls. PLoS One 7, e37062 (2012).

29. O. Aziz, J. Klenk, L. Schwickert, L. Chiari, C. Becker, E. J. Park, G. Mori, S. N. Robinovitch, Validation of accuracy of SVM-based fall detection system using real-world fall and non-fall datasets. PLoS One 12, e0180318 (2017).

30. J. Klenk, C. Becker, F. Lieken, S. Nicolai, W. Maetzler, W. Alt, W. Zijlstra, J. M. Hausdorff, R. C. van Lummel, L. Chiari, U. Lindemann, Comparison of acceleration signals of simulated and real-world backward falls. Med Eng Phys 33, 368–373 (2011).

31. J. Silva, I. Sousa, J. Cardoso, “Transfer learning approach for fall detection with the FARSEEING real-world dataset and simulated falls” in 2018 40th Annual International Conference of the IEEE Engineering in Medicine and Biology Society (EMBC) (IEEE, 2018), pp. 3509–3512.

32. Wolk Airbag. Wolk Airbag Safety System. Wolk Airbag, Available at: https://wolkairbag.com. Accessed February, 2025.

33. Scikit-learn Developers, Scikit-learn: Machine Learning in Python. Available at: https://scikit-learn.org/stable/ Accessed February, 2025.

34. J. R. Crenshaw, K. A. Bernhardt, S. J. Achenbach, E. J. Atkinson, S. Khosla, K. R. Kaufman, S. Amin, The circumstances, orientations, and impact locations of falls in community-dwelling older women. Arch Gerontol Geriatr 73, 240–247 (2017).

35. J. Lo, J. A. Ashton-Miller, Effect of Upper and Lower Extremity Control Strategies on Predicted Injury Risk During Simulated Forward Falls: A Study in Healthy Young Adults. J Biomech Eng 130 (2008).

36. J. Klenk, L. Schwickert, L. Palmerini, S. Mellone, A. Bourke, E. A. F. Ihlen, N. Kerse, K. Hauer, M. Pijnappels, M. Synofzik, K. Srulijes, W. Maetzler, J. L. Helbostad, W. Zijlstra, K. Aminian, C. Todd, L. Chiari, C. Becker, The FARSEEING real-world fall repository: a large-scale collaborative database to collect and share sensor signals from real-world falls. European Review of Aging and Physical Activity 13, 8 (2016).

37. R. W. Broadley, J. Klenk, S. B. Thies, L. P. J. Kenney, M. H. Granat, Methods for the Real-World Evaluation of Fall Detection Technology: A Scoping Review. Sensors 18, 2060 (2018).

38. H. Gaßner, P. Sanders, A. Dietrich, F. Marxreiter, B. M. Eskofier, J. Winkler, J. Klucken, Clinical Relevance of Standardized Mobile Gait Tests. Reliability Analysis Between Gait Recordings at Hospital and Home in Parkinson’s Disease: A Pilot Study. J Parkinsons Dis 10, 1763–1773 (2020).

39. C. Bottari, É. Dutil, C. Dassa, C. Rainville, Choosing the most appropriate environment to evaluate independence in everyday activities: Home or clinic? Aust Occup Ther J 53, 98– 106 (2006).

40. A. K. Bourke, P. van de Ven, M. Gamble, R. O’Connor, K. Murphy, E. Bogan, E. McQuade, P. Finucane, G. ÓLaighin, J. Nelson, Evaluation of waist-mounted tri-axial accelerometer based fall-detection algorithms during scripted and continuous unscripted activities. J Biomech 43, 3051–3057 (2010).

41. L. Palmerini, J. Klenk, C. Becker, L. Chiari, Accelerometer-Based Fall Detection Using Machine Learning: Training and Testing on Real-World Falls. Sensors 20, 6479 (2020).

42. O. Aziz, C. M. Russell, E. J. Park, S. N. Robinovitch, “The effect of window size and lead time on pre-impact fall detection accuracy using support vector machine analysis of waist mounted inertial sensor data” in 2014 36th Annual International Conference of the IEEE Engineering in Medicine and Biology Society (IEEE, 2014), pp. 30–33.

43. C. Mosquera-Lopez, E. Wan, M. Shastry, J. Folsom, J. Leitschuh, J. Condon, U. Rajhbeharrysingh, A. Hildebrand, M. Cameron, P. G. Jacobs, Automated Detection of Real-World Falls: Modeled From People With Multiple Sclerosis. IEEE J Biomed Health Inform 25, 1975–1984 (2021).

44. E. Stack, Falls are unintentional: Studying simulations is a waste of faking time. J Rehabil Assist Technol Eng 4 (2017).

45. J. K. Lee, S. N. Robinovitch, E. J. Park, Inertial Sensing-Based Pre-Impact Detection of Falls Involving Near-Fall Scenarios. IEEE Transactions on Neural Systems and Rehabilitation Engineering 23, 258–266 (2015).

46. Y. Harari, N. Shawen, C. K. Mummidisetty, M. V. Albert, K. P. Kording, A. Jayaraman, A smartphone-based online system for fall detection with alert notifications and contextual information of real-life falls. J Neuroeng Rehabil 18, 124 (2021).

47. C. Medrano, R. Igual, I. Plaza, M. Castro, Detecting Falls as Novelties in Acceleration Patterns Acquired with Smartphones. PLoS One 9, e94811 (2014).

48. M. Kangas, I. Vikman, L. Nyberg, R. Korpelainen, J. Lindblom, T. Jämsä, Comparison of real-life accidental falls in older people with experimental falls in middle-aged test subjects. Gait Posture 35, 500–505 (2012).

49. M. Kangas, A. Konttila, P. Lindgren, I. Winblad, T. Jämsä, Comparison of low-complexity fall detection algorithms for body attached accelerometers. Gait Posture 28, 285–291 (2008).

50. T. E. Lockhart, R. Soangra, H. Yoon, T. Wu, C. W. Frames, R. Weaver, K. A. Roberto, Prediction of fall risk among community-dwelling older adults using a wearable system. Sci Rep 11, 20976 (2021).

51. B. Nemeth, M. van der Kaaij, R. Nelissen, J.-K. van Wijnen, K. Drost, G. J. Blauw, Prevention of hip fractures in older adults residing in long-term care facilities with a hip airbag: a retrospective pilot study. BMC Geriatr 22, 547 (2022).

52. A. Choi, T.-H. Kim, Y. O, S. Jeong, K. Kim, H. Kim, J.-H. Mun, Deep learning-based near-fall detection algorithm for fall risk monitoring system using a single inertial measurement unit. IEEE Trans. Neural Syst. Rehabil. Eng. 30, 2385–2394 (2022).

53. J. Silva, et al., Automated development of custom fall detectors: Position, model, and rate impact in performance. IEEE Sens. J. 20, 5465–5472 (2020).

54. W. Saadeh, S. A. Butt, M. A. B. Altaf, A patient-specific single sensor IoT-based wearable fall prediction and detection system. IEEE Trans. Neural Syst. Rehabil. Eng. 27, 995–1003 (2019).

55. J.-S. Lee, H.-H. Tseng, Development of an enhanced threshold-based fall detection system using smartphones with built-in accelerometers. IEEE Sens. J. 19, 8293–8302 (2019).

56. A. Shahzad, K. Kim, FallDroid: An automated smartphone-based fall detection system using multiple kernel learning. IEEE Trans. Ind. Inform. 15, 35–44 (2018).

57. A. Altmann, L. Toloşi, O. Sander, T. Lengauer, Permutation importance: a corrected feature importance measure. Bioinformatics 26, 1340–1347 (2010).

58. N. Segev, M. Harel, S. Mannor, K. Crammer, R. El-Yaniv, Learn on Source, Refine on Target: A Model Transfer Learning Framework with Random Forests. IEEE Trans Pattern Anal Mach Intell 39, 1811–1824 (2017).

59. Luke3D. Transfer Random Forest. GitHub 2024. Available at: https://github.com/Luke3D/TransferRandomForest Accessed February, 2025.

60. C. Chen, A. Liaw, L. Breiman, others, Using random forest to learn imbalanced data. *University of California*, Berkeley 110, 24 (2004).

