## Supplementary figures and images for "From Lab to Life: Enhancing Wearable Airbag Fall Detection Performance with Minimal Real-World Data"

### Supplemental Figure 1

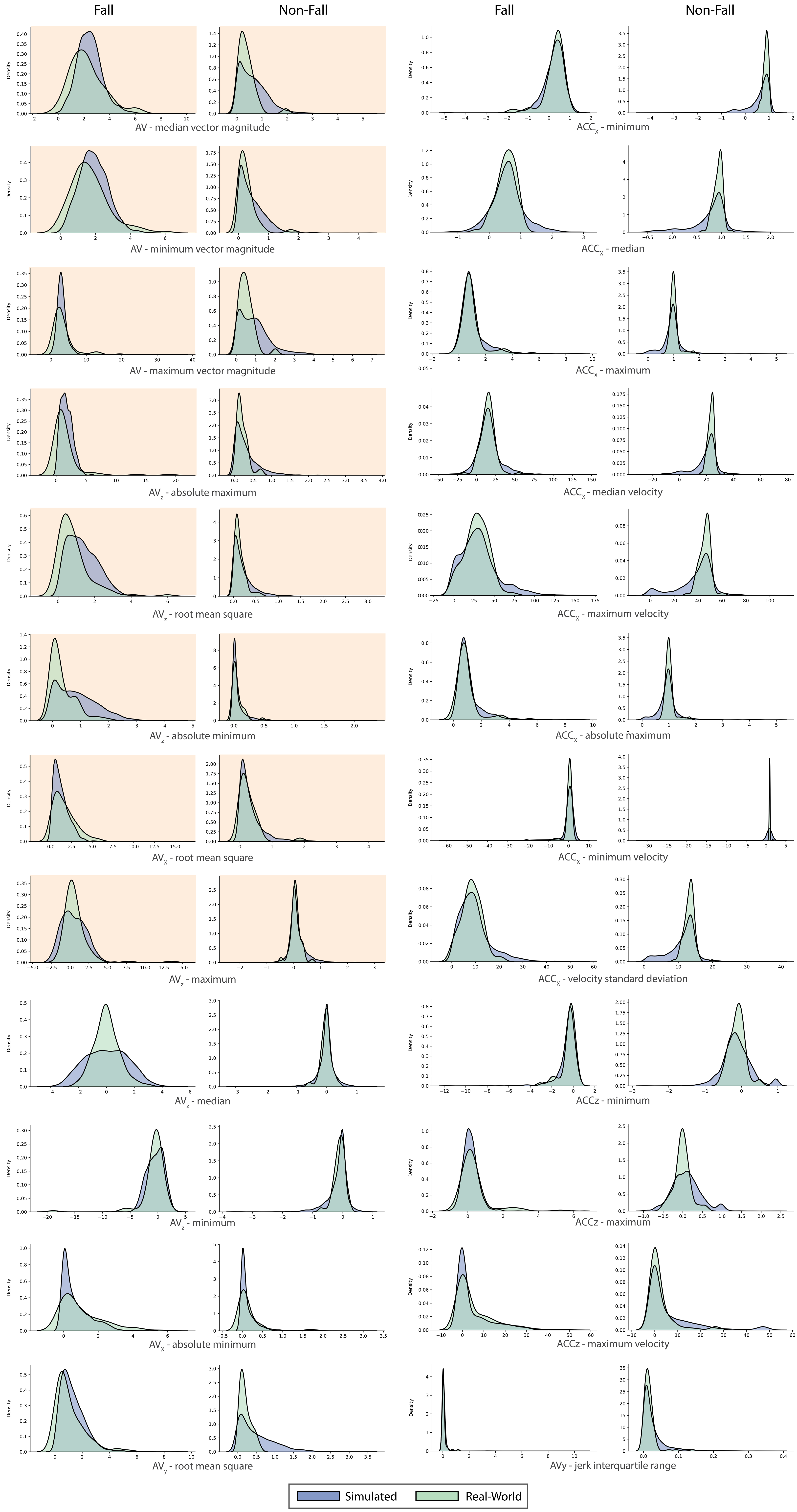

### Supplemental Figure 2

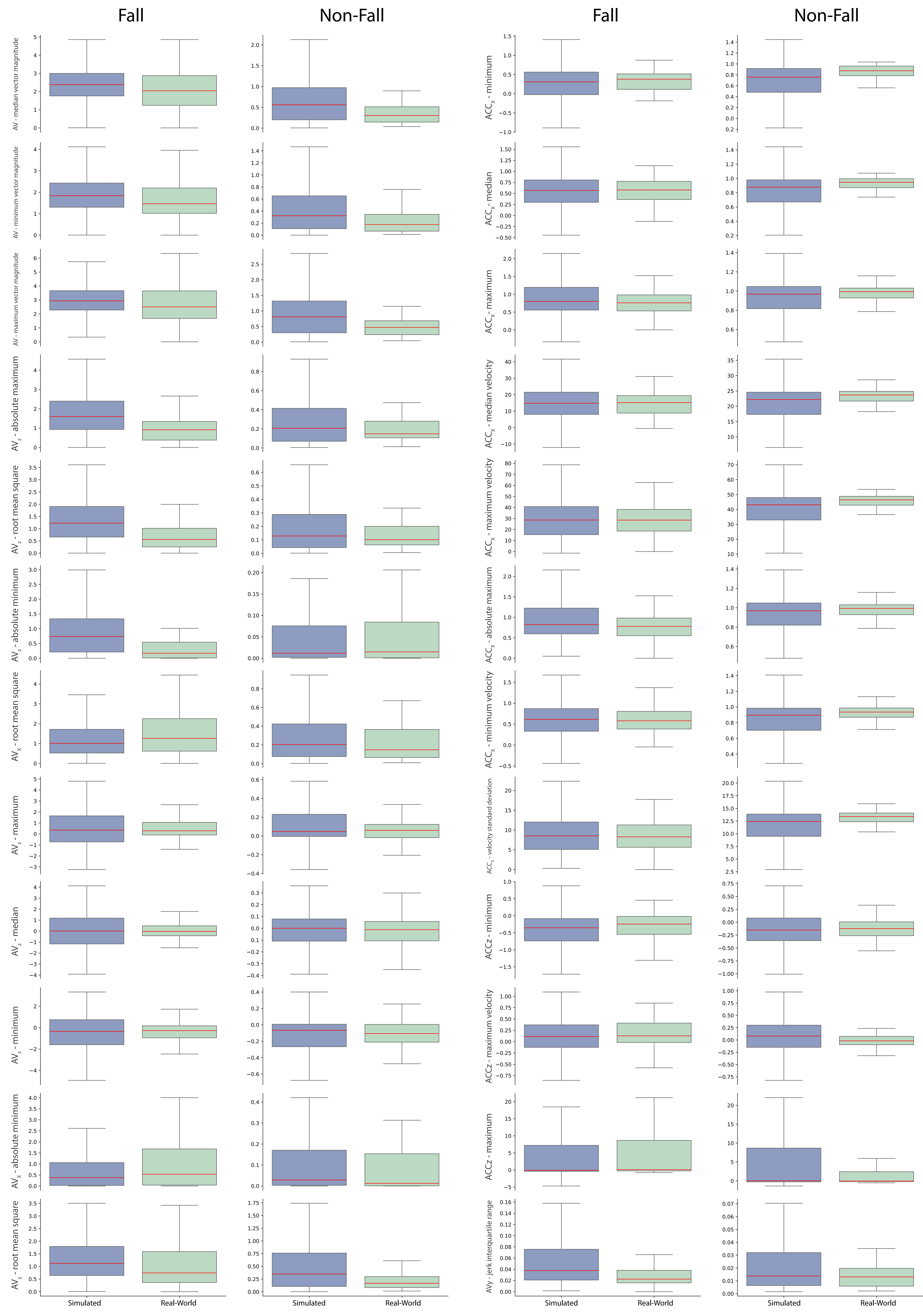
